# Periodic COVID-19 Testing in Emergency Department Staff

**DOI:** 10.1101/2020.04.28.20084053

**Authors:** Yuemei Zhang, Sheng-Ru Cheng

**Affiliations:** Department of Anesthesiology and Pain Medicine, University of Washington, Seattle, WA, USA; Dept. of Electrical and Computer Engineering, University of Illinois at Urbana-Champaign, Urbana, IL, USA

## Abstract

**Background:** As the number of COVID-19 cases in the US continues to rise and hospitals are experiencing personal protective equipment (PPE) shortages, healthcare workers have been disproportionately affected by COVID-19 infection. Since COVID-19 testing is now available, some have raised the question of whether we should be routinely testing asymptomatic healthcare workers.

**Methods:** Using publicly available data on COVID-19 infections and emergency department visits, as well as internal hospital staffing information, we generated a mathematical model to predict the impact of periodic COVID-19 testing in asymptomatic members of the emergency department staff in regions affected by COVID-19 infection. We calculated various transmission constants based on the Diamond Princess cruise ship data, used a logistic model to calculate new infections, and we created a Markov model according to average COVID-19 incubation time.

**Results:** Our model predicts that after 30 days, with a transmission constant of 1.219e-4 new infections per person^2^, weekly COVID-19 testing of healthcare workers (HCW) would reduce new HCW and patient infections by 5.1% and bi-weekly testing would reduce both by 2.3%. At a transmission constant of 3.660e-4 new infections per person,^2^ weekly testing would reduce infections by 21.1% and bi-weekly testing would reduce infections by 9.7–9.8%. For a lower transmission constant of 4.067e-5 new infections per person^2^, weekly and biweekly HCW testing would result in a 1.54% and 0.7% reduction in infections respectively.

**Conclusion:** Periodic COVID-19 testing for emergency department staff in regions that are heavily-affected by COVID-19 and/or facing resource constraints may reduce COVID-19 transmission significantly among healthcare workers and previously-uninfected patients.

## Introduction

Although it originated as a small cluster of cases restricted to Wuhan, China in Nov. and Dec. of 2019, COVID-19 rapidly spread across the globe and officially reached “pandemic” status on March 11, 2020.^1^ In the United States, the number of confirmed cases spiked from just 1 case in Jan. 20, 2020 to 612,576 confirmed positives and 29,798 deaths as of April 14, 2020.^2^ Washington state, the location of the first American case, has had 10,538 COVID19+ patients as of April 14, 2020.^3^ Given its rapid spread and 3.4% mortality rate,^4^ countries like Italy and China have been forced to ration limited healthcare resources, and there are concerns that the US may need to do so as well.^5^ Person-to-person transmission by asymptomatic individuals and pre-symptomatic individuals during the up-to-14 day incubation period^6^ may play a significant role in this pandemic.^7–10^

Because of the higher risk of exposure to COVID19+ patients and shortages in personal protective equipment (PPE) both in the US and in other countries,^11–13^ healthcare workers have been disproportionately affected by COVID-19 infection.^14–16^

The goal of this study is to provide a quantitative analysis and model for predicting the impact of periodic COVID-19 testing for all emergency room staff as a possible alternate strategy to mitigate disease transmission in the healthcare setting, since PPE supplies are limited.

## Methods

### Data sourcing

In order to model a hospital emergency department and a moderately affected patient population, we chose to base our model on Harborview Medical Center (HMC) in King County, WA, since we had access to its emergency department (ED) staffing information. Because Harborview is one of many hospitals within the region, for the sake of simplicity, we are assuming that HMC’s entire patient population essentially lives in King County, WA.

In order to estimate the number of daily ED visits, we used the publicly available UW Medicine Annual Financial Report for the Board of Regents meeting, which reported 57,516 ED visits to HMC during fiscal year 2018.^17^ Next, we divided this. number by 365 days / year, since medical emergencies happen daily regardless of holidays, to estimate average daily ED visits. Although it is possible that the rate of ED visits has changed due to COVID-19 symptoms or due to socio-behavioral changes resulting from the COVID-19 pandemic and/or public policies related to it, this number is not currently available to us.

The HMC emergency department currently employs 59 emergency medicine (EM) faculty physicians, 48 EM resident physicians, and 200 full-time equivalent registered nurses (RNs) and medical assistants (MAs). Together, these sum to the equivalent of 307 full-time healthcare workers in the ED.

**Fig 1.**
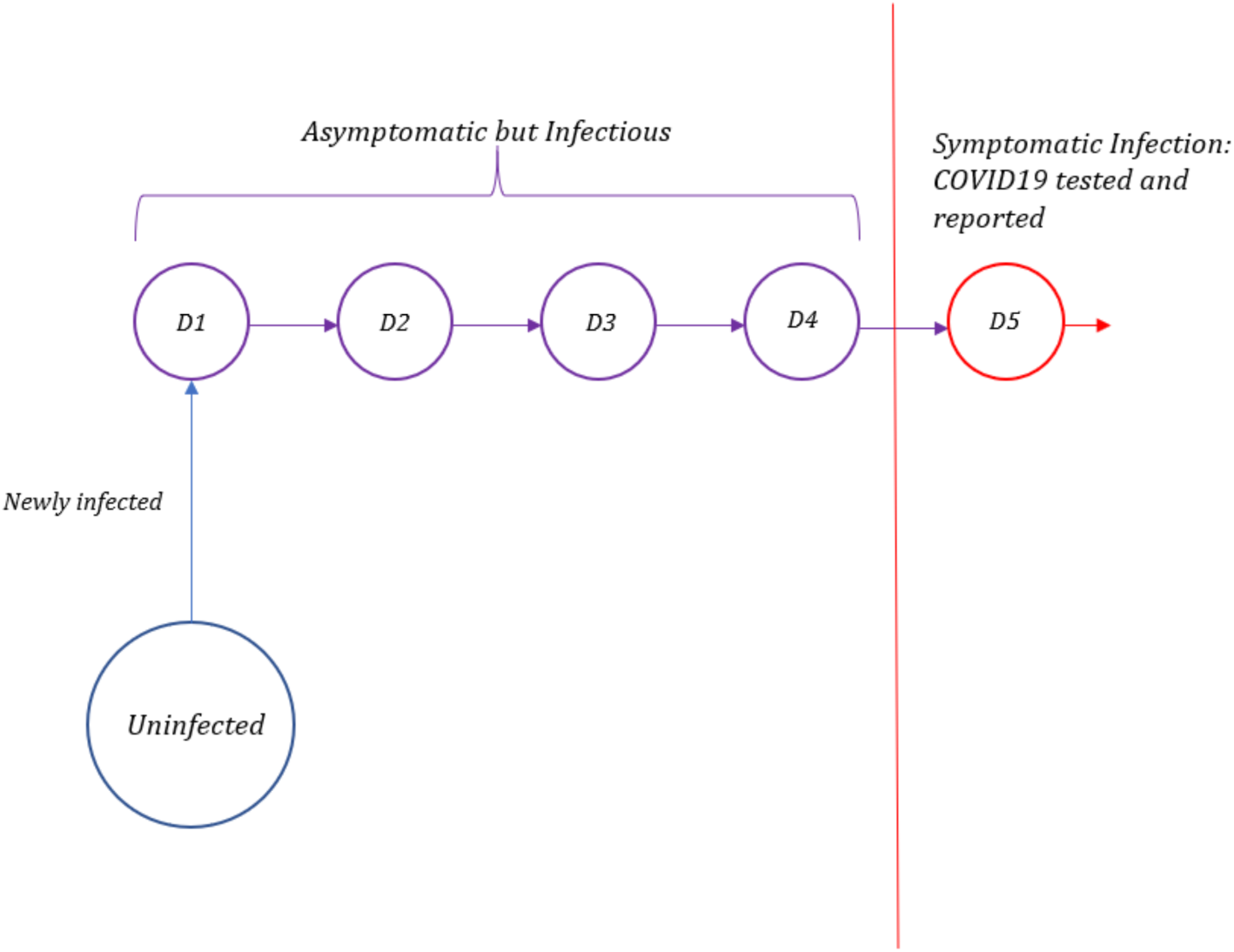
Timeline of Infection for Confirmed COVID-19 Cases. After infection, the individual can transmit the infection to others but does not become symptomatic until day 5.

### Initial conditions

Since our model is intended to be generalizable to any hospital in the United States, we did not apply HMC-specific policies to our model and instead maintained the same constraints that many other US hospitals have.

Due to the incubation period of the virus, coupled with the current resource limitations in the US, COVID-19 testing is often not performed until symptoms become evident. Furthermore, due to laboratory processing times, COVID-19 test results may not be available until patients have left the emergency department. To estimate the asymptomatic infected population, we looked at the number of newly-confirmed COVID-19 cases on each date and retroactively calculated the daily number of individuals that would have been in the pre-symptomatic incubation phase. Based on recently published studies, the average incubation period of COVID-19 is around 5 to 6 days.^18–20^ For this model, we used the shorter incubation period of 5 days, meaning that symptoms begin on day 5. This means that, for any time t, the number of asymptomatic but infected individuals can be estimated using the sum of new infections that were confirmed on t + 1 to t + 4 as follows:

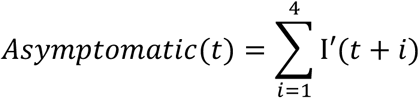

‘In other words, if someone is symptomatic and confirmed to be COVID19+ on any of the days between t+1 to t+4, then s/he was infected but asymptomatic on day t. Using data for King County, WA through April 5, 2020, we calculated 671 asymptomatic cases on April 1, 2020 in King County.

In order to determine the total number of infected individuals in King County on any given date, we then added the number of publicly reported confirmed infections on April 1, 2020 with the asymptomatically-infected population that we calculated and subtracted the number of COVID-19 deaths. To determine the total uninfected population, we subtracted the infected population and the number of COVID-19 deaths from King County’s estimated 2019 population of 2,252,782.^21^ Subsequently, to determine the proportion of the living population that was infected and uninfected respectively, we divided the total infected population and the total uninfected population by the total living population.

We assume that since the majority of patients and HCWs reside locally, their infection statuses would initially also be representative of that of the general population. Thus we multiplied our proportions with 157.56 total emergency department (ED) patients per day and 307 total HCW in the ED to arrive at the initial values of 0.21 infected patients per day, 157.36 uninfected patients per day, 0.41 infected HCW, and 306.57 uninfected HCW. Infected HCW were further subdivided into groups based on how long they had been infected. Because asymptomatic COVID19+ individuals would remain in the workforce, we included infected HCW in the healthcare workforce for days 1–4 of their infections (during which time they could also infect other HCW and patients), and then removed them from the workforce once they reached day 5 and displayed symptoms. For our initial conditions, we divided infected HCW evenly into 4 groups for HCW on day 1 of infection (D1), day 2 of infection (D2), day 3 of infection (D3) day 4 of infection (D4).

### Transmission rate

To investigate the number of preventable infections of healthcare workers from asymptomatic infected patients, we used a simple logistic model of transmission:

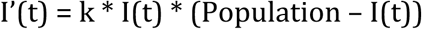

In this equation, k is the transmission constant, I’(t) is the rate of change of infected population, and I(t) represents total infected population, including the asymptomatic infected population. Since I’(t) is the rate of change of the infected population, it can be observed that the total infected population at a discrete time t + 1 is calculated as

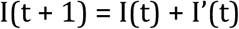

To calculate the transmission constant, we rearrange the previous equations to the following

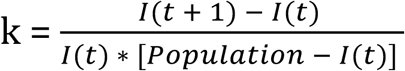

Since we are interested in the total infection spread, data for some known infected population, both symptomatic and asymptomatic, is required. For this, we used data extracted from the Diamond Princess cruise ship,^22^ since the close quarters approximate the clinical setting. Due to the isolated nature of the ship, health officials were able to test everyone onboard the cruise ship, even if there are no symptoms evident. Using the data at hand and the equation above, we can readily determine the transmission constant by dividing the number of new cases at time t + 1 (with time measured in days) by the product of infected population at time t and the uninfected population at time t, which we calculated to be an average of k = 1.219e-4 new infections per person^2^.

While we know the transmission rate for healthcare workers is likely different, we do not know whether it is higher or lower, due to both more intimate and close contact with patients than typical interactions on a cruise ship, and due to questions of PPE usage. Furthermore, the transmission rate likely varies by department and institution as well. Since we do not have an accurate transmission rate for the resource-limited clinical environment, we decided to model several different scenarios using 3 times the transmission constant (3.660e-4 new infections per person^2^) and one-third the transmission constant (4.067e-5 new infections per person^2^) calculated from the Diamond Princess cruise ship scenario.

To calculate the number of patient-to-HCW infections, HCW-to-patient infections, and HCW-to-HCW infections occurring in the ED, we adapted the logistic model to

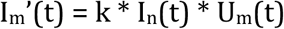

where I_m_’(t) refers to the new infections of a population *m*, k is the transmission constant, I_n_(t) refers to asymptomatically infected individuals of the group *n* transmitting the virus, and U_m_(t) refers to uninfected individuals of the group *m* that is being newly infected. For instance, if I_m_’(t) represents new HCW-to-patient infections, then I_n_(t) would represent asymptomatically infected HCW, and U_m_(t) would represent uninfected patients presenting to the emergency department. These calculations would be repeated for every day in our model.

Assuming adequate inpatient beds, patients leave the ED each day, whether that means they were admitted to the hospital or are leaving the institution, and a new batch of patients with characteristics representative of the general population would arrive each day. Therefore, the starting numbers of uninfected patients and infected patients that we used for our calculations stayed constant. In reality, the number of COVID19+ patients presenting to the ED may be disproportionately higher than in the general population, since completely healthy individuals without any acute illness or injury would not visit the ED.

On the other hand, since HCW were unlikely to have significant changes in their employment in the time period we were modeling, we designed a Markov chain to track their infection timelines. New HCW infections comprised the D1 group for the following day, and HCW in D1 would get changed to D2 the following day, HCW in D2 would get changed to D3 the following day, so on and so forth.

### Periodic Testing

To simulate periodic COVID-19 testing of all HCW, we used the simplified case of 100% sensitivity for COVID-19 testing. In reality, testing sensitivity is likely to be lower and may vary based on how testing or sample collection is performed, and our model can be adapted for other levels of sensitivity. Currently, there is insufficient data on testing to have information on sensitivity. On any given day that all HCW are tested, we would manually remove (sensitivity)*(number of infected HCW on each day) from each category. With 100% sensitivity, this would mean that all infected HCW on the testing day. For weekly testing, we started this manual elimination process on day 6, and then repeated this process every 7 days. For biweekly testing, we started the manual elimination process on day 13, and then repeated this process every 14 days.

**Figure 2.**
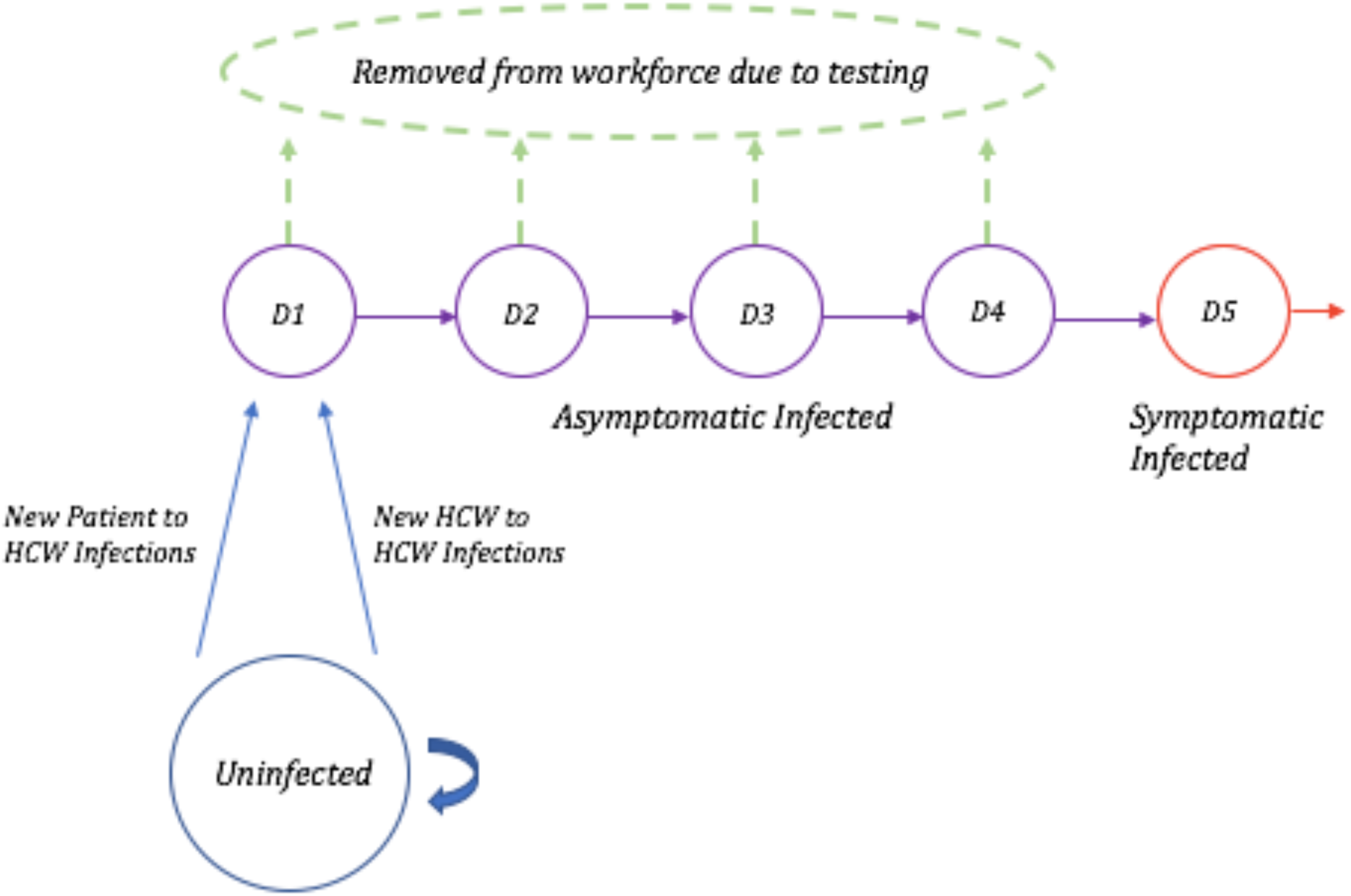
Markov Chain for Healthcare Workers. Healthcare workers (HCW) who are uninfected on any given day can either stay uninfected or become newly infected (blue), at which point they would proceed to day 1 of infection the next day. Individuals who are infected will proceed to the next day of infection with each passing day. Infected HCW are asymptomatic on days 1–4 (purple). On day 5 of infection, infected individuals begin showing symptoms, at which point they are removed from this workforce (red). With COVID-19 testing, On days with COVID-19 testing, asymptomatic infected HCW who test positive may also be removed from the healthcare workforce (green).

## Results

Our model predicts that over the course of 30 days, 4727 patients visited the emergency department in one hospital. At the baseline transmission constant of 1.219e-4 new infections per person^2^, without routine COVID-19 testing of HCW, 0.318 HCW infections and 0.163 new patient infections would occur in one hospital emergency department after these 30 days. If COVID-19 testing of HCW occurred every 7 days, then 0.302 HCW infections and 0.155 new patient infections would occur, which is a 5.1% reduction in both HCW and new patient infections. If COVID-19 testing of HCW occurred every 14 days, then 0.311 HCW infections and 0.160 new patient infections would occur, which is a 2.3% reduction in both HCW and new patient infections.

**Table 1.** Patient Infections with and without Periodic COVID-19 Testing for HCW. Predicted numbers of new patient infections with various transmission rates and COVID-19 testing frequencies for healthcare workers (HCW) after 30 days within 1 hospital emergency department. Percentages in parentheses represent the decrease in number of infections at each transmission rate with weekly (every 7 days) or biweekly (every 14 days) testing compared to the number of infections if HCW were not routinely tested.

| Patient Infections with and without Periodic COVID-19 Testing for HCW |  |  |  |
| --- | --- | --- | --- |
| Transmission Rate | No Testing | Weekly Testing | Biweekly Testing |
| $1.219\text{e-}4$ new infections/person<sup>2</sup> | 0.163 | 0.155 (5.1%) | 0.160 (2.3%) |
| $3.660\text{e-}4$ new infections/person<sup>2</sup> | 0.720 | 0.568 (21.1%) | 0.650 (9.8%) |
| $4.067\text{e-}5$ new infections/person<sup>2</sup> | 0.0491 | 0.0484 (1.54%) | 0.0488 (0.7%) |

With a transmission constant of 3.660e-4 new infections per person^2^, without routine COVID-19 testing of HCW, 1.40 HCW infections and 0.720 new patient infections would occur in one hospital emergency department. If COVID19 testing of HCW occurred every 7 days, then 1.10 HCW infections and 0.568 new patient infections would occur, which is a 21.1% reduction in both HCW and new patient infections. If COVID19 testing of HCW occurred every 14 days, then 1.26 HCW infections and 0.650 new patient infections would occur, which is a 9.7% reduction in HCW infections and 9.8% reduction in new patient infections.

**Table 2.** HCW Infections with and without Periodic COVID-19 Testing for HCW. Predicted numbers of new HCW infections with various transmission rates and COVID-19 testing frequencies for healthcare workers after 30 days within 1 hospital emergency department. Percentages in parentheses represent the decrease in number of infections at each transmission rate with weekly (every 7 days) or biweekly (every 14 days) testing compared to the number of infections if HCW were not routinely tested.

| HCW Infections with and without Periodic COVID-19 Testing for HCW |  |  |  |
| --- | --- | --- | --- |
| Transmission Rate | No Testing | Weekly Testing | Biweekly Testing |
| 1.219e-4 new infections/person<sup>2</sup> | 0.318 | 0.302 (5.1%) | 0.311 (2.3%) |
| 3.660e-4 new infections/person<sup>2</sup> | 1.40 | 1.10 (21.2%) | 1.26 (9.7%) |
| 4.067e-5 new infections/person<sup>2</sup> | 0.0957 | 0.0943 (1.54%) | 0.0951 (0.7%) |

For a lower transmission constant of 4.067e-5 new infections per person^2^, 0.0957 HCW infections and 0.0491 new patient infections would occur in one hospital emergency department without routine COVID-19 testing of HCW. If COVID19 testing of HCW occurred every 7 days, then 0.0943 HCW infections and 0.0484 new patient infections would occur, which is a 1.54% reduction in both HCW and new patient infections. If COVID19 testing of HCW occurred every 14 days, then 0.0951 HCW infections and 0.0488 new patient infections would occur, which is a 0.7% reduction in HCW infections and new patient infections.

## Discussion

This model shows that within a single hospital emergency department, periodic COVID-19 testing among healthcare workers would reduce the rate of COVID-19 infections among emergency department personnel and reduce the rate of new COVID-19 infections acquired by patients in the ED. As expected, the impact of periodic HCW testing varied with the transmission rate of COVID-19, showing greater benefit when COVID-19 transmission rates were higher.

Our model uses the COVID-19 prevalence for King County, WA, an area which is not as heavily-affected by COVID-19 as many other places in this country, for the disease prevalence in patients arriving to the ED. A higher COVID-19 prevalence for the patient population may result in higher patient-to-HCW disease transmission rates, in which case, periodic HCW testing would be more beneficial.

A limitation of our model is that we do not know the actual transmission rate in various hospital emergency departments, and transmission rates may vary widely between hospitals based on PPE supply, type of interactions with patients and severity of illness (which also impacts the types of procedures and therapies involved), and other factors. Additionally, the transmission rate may be different for different types of healthcare workers: for instance, those who perform aerosolizing procedures like intubation may experience a higher rate of transmission.

By changing the initial parameters, this model can be adapted for different ED visit rates, different ED staffing numbers, different levels of infection prevalence, different transmission constants, and different levels of testing sensitivity. Lower levels of testing sensitivity will lead to decreased utility in periodic HCW testing. In addition, our analysis was performed with the population characteristics of a county that is moderately affected by COVID-19. Currently, there are many regions of the country with a much higher COVID-19 prevalence, which would lead to a greater potential benefit from periodic HCW testing to prevent HCW infections.

Due to the current state of COVID-19 testing, US statistics on confirmed COVID-19 cases may not be the most reliable, either. Per CDC guidelines that were last updated March 24,2020, laboratory testing for COVID-19 is only indicated for individuals who are not healthcare workers nor first responders if they have symptoms that are consistent with COVID-19.^23^ However, many COVID19+ individuals may be asymptomatic or only have mild symptoms.^24^ In addition, COVID-19 testing shortages may make the US statistics on COVID-19 cases less reliable.^25^ Therefore, the numbers for COVID-19 incidence and prevalence used in our model, which are based on official reports, may be erroneously low.

Of note, our model only includes emergency department staff in our numbers, but healthcare workers from other specialties and departments also see patients in the ED. For instance, in many hospitals, non-emergency medicine physicians will see inpatient admissions in the ED, specialists may be consulted to see patients in the ED, and surgeons and anesthesiologists participate in trauma resuscitations. Additionally, at some teaching hospitals, resident physicians in specialties outside of emergency medicine will also have emergency medicine rotations.

Given the uncertainty and unavailable data regarding COVID-19, some of the numbers and factual assumptions in this model may be incorrect, which could affect the model’s predictions. To simplify calculations, this model assumes that COVID-19 infections are spread homogeneously throughout the state, that healthcare workers freely interact with patients and all other healthcare workers, and does not take into account individual variation in incubation times. Ultimately, this model is intended to be a tool and an approximation, and it can be adapted to different healthcare settings or regions by changing the starting conditions.

## Data Availability

The data that support the findings of this study are openly available at the links provided.

https://github.com/CSSEGISandData/COVID-19

https://www.doh.wa.gov/Emergencies/Coronavirus

https://s3-us-west-2.amazonaws.com/uw-s3-cdn/wp-content/uploads/sites/12/2019/02/06104924/2019-02-B-6.pdf

https://www.census.gov/quickfacts/fact/table/kingcountywashington,WA/PST045218

https://doi.org/10.1016/S1473-3099(20)30120-1

